# Pandemic-related changes in postpartum depression and anxiety among breastfeeding mothers: a systematic review and meta-analysis

**DOI:** 10.64898/2026.05.18.26353483

**Authors:** Jinyue Yu, Matthew McCann, Michael Clesham, Mary Fewtrell

## Abstract

**Background:** The COVID-19 pandemic caused major disruptions to maternity care, breastfeeding support, and social networks. These changes may have increased the risk of postpartum depression, anxiety, and stress among breastfeeding mothers, a population that has been underrepresented in previous reviews. This systematic review and meta-analysis aimed to compare maternal mental health outcomes among breastfeeding mothers before and during the COVID-19 pandemic.

**Methods:** We searched MEDLINE, EMBASE, AMED, Web of Science, WanFang Data, MedRxiv, WHO COVID-19 databases, and grey literature from database inception to December 2023. Eligible studies compared mental health outcomes in breastfeeding mothers before and during the COVID-19 pandemic using validated assessment tools, including the Edinburgh Postnatal Depression Scale (EPDS), Generalized Anxiety Disorder Scale (GAD-7), State-Trait Anxiety Inventory (STAI), or Perceived Stress Scale (PSS). Studies with fewer than 10 participants per group were excluded. Two reviewers independently screened studies, extracted data, and assessed risk of bias using the Joanna Briggs Institute checklist or Newcastle-Ottawa Scale, depending on study design. Random-effects meta-analysis was performed when at least two studies reported comparable outcomes.

**Results:** Twenty-three studies involving breastfeeding mothers from 15 countries were included. Meta-analysis showed significantly higher depressive symptoms during the pandemic compared with the pre-pandemic period, measured by EPDS (standardized mean difference [SMD] = 0.21, 95% confidence interval [CI] 0.14 to 0.29). Maternal anxiety measured by GAD-7 was also significantly higher during the pandemic (SMD = 0.27, 95% CI 0.13 to 0.41). Findings for perceived stress were mixed across studies and could not be pooled because of heterogeneity in reporting methods. Limited evidence suggested that mother-infant bonding did not substantially decline during the pandemic despite increased maternal psychological distress.

**Conclusions:** Breastfeeding mothers experienced increased postpartum depression and anxiety symptoms during the COVID-19 pandemic. These findings highlight the importance of maintaining breastfeeding support services, ensuring access to maternal mental health screening, and developing flexible models of postpartum care during future public health emergencies.

**PROSPERO registration:** CRD42022354670.

## Introduction

The 2019 coronavirus disease (COVID-19) pandemic was a global crisis that affected every aspect of life [1,2]. In an effort to minimize the transmission of the virus, various preventive measures such as “stay-at-home”, “lockdown” measures and adherence to social distancing guidelines were implemented globally [3]. Whilst such approaches were effective in preventing the transmission of the virus, there were multiple side effects including consequences for the mental health of the general population, particularly amongst vulnerable groups. Mothers who were feeding their infants during the pandemic face heightened challenges exacerbated by disruption to routine healthcare visits and lactation support services, which posed significant obstacles to breastfeeding initiation and continuation [4,5]. Postpartum women also suffered anxiety related to loss of employment, reduced salary [3,6], and fears of transmitting the virus to their newborn. Research suggests that depression and distress among postpartum women may have increased during this period [7-9].

Given the important role of infant feeding in maternal-infant bonding and on infant health outcomes, it is important to explore the mental health implications of these challenges for mothers who were feeding their infants during the pandemic. However, existing systematic reviews in this area focused on pregnant and perinatal women, with a lack of comprehensive systematic reviews focusing on breastfeeding women. While individual studies have offered valuable insights, the lack of synthesis and consensus limits our understanding of the full scope and implications of maternal mental health in the context of the pandemic.

To address this gap, we conducted a systematic review to investigate the impact of Covid-19 pandemic on maternal mental health status among lactating women. In the present study, we focused on studies that assessed maternal depression and anxiety using validated questionnaires such as the Edinburgh Postnatal Depression Scale (EPDS) in groups of women before and during the period of pandemic restrictions. By summarising studies that compare mothers’ mental health status across these time points, we aim to provide greater understanding of the impact of the pandemic restrictions among countries, and to discuss the potential determinants and moderators of psychological distress that can be affected by different culture and social norms. Whilst the pandemic is over, these findings may inform practice and policy during future emergency situations.

## Methods

### Study design and search strategy

We conducted a systematic review of all studies evaluating depressive, anxiety, and stress symptoms of breastfeeding women during the Covid-19 pandemic (published after (January 2020). The design, interpretation of data, drafting and revisions followed the PRISMA guideline. The literature search was performed in Ovid MEDLINE (1946-2022 August 09), EMBASE (1980-2022 week 31), AMED (1985-August 2022), Web of Science, MedRxiv, WHO’s Global Research on Coronavirus Disease (COVID-19) WanFang Data, and the reference lists of the included papers. The search strategy was conducted using the combination of the following Medical Subject Heading (MeSH) terms and any relevant keywords: (“COVID-19” OR “2019-nCoV” OR “novel coronavirus” OR SARS-CoV-2 OR “coronavirus 2”) AND (breastfeeding OR breast feed OR “infant feeding” OR “lactation” OR “postnatal” “postpartum”) AND (“anxiety” OR “depression” OR “depressive” OR “stress” OR “distress” OR “coping”). Two independent researchers (JY and MM) completed the title, abstract, and full-text review from August 2022 to May 2023). Any disagreement was resolved by consensus and, whenever necessary, a third external collaborator was consulted (MF). The protocol for the systematic review was registered on PROSPERO (CRD42022354670).

### Inclusion and exclusion criteria

Inclusion criteria for the registered systematic review were: 1) study participants were postnatal women over 18 years of age and had breast-fed their infant during the Covid-19 pandemic; 2) infants were under 18 months during the Covid-19 pandemic; 3) infants were generally healthy; 4) study of any design that provided quantitative data on maternal mental health or infant feeding outcomes. Exclusion criteria were as follows; 1) participants were women during pregnancy without postpartum follow-up data; 2) infants had any condition that might affect infant feeding; 3) case-report, qualitative study, any kind of review, study protocol, and studies with no full text available.

Since the present review specifically focused on differences in maternal mental health status before and during the pandemic period, we only included studies that reported the maternal mental health status in groups of women before and during the pandemic using a validated questionnaire, such as EPDS, Generalized Anxiety Disorder Scale (GAD-7), State-Trait Anxiety Inventory (STAI), Beck Anxiety Inventory (BAI), and Perceived Stress Scale (PSS).

### Data extraction

Data extraction was conducted by MC, MF, JY, and checked by each other. Any disagreement was resolved by consensus. The following data were extracted: first author’s name, year of publication, study design, study setting, study timeline, number of participants, maternal and infant’s characteristics, outcomes and measurements, main results, and mean and standard deviation (SD) of assessment tool scores where applicable.

### Data synthesis

Extracted data were synthesised using both narrative and quantitative approaches. We first conducted a descriptive synthesis to summarise the key characteristics of each included study, including study design, country, sample size, maternal and infant demographics, mental health assessment tools used, and relevant outcomes. All information was extracted into tables to identify variations in study settings and methodologies.

### Meta-analysis

When there were two or more studies using the same valid instrument to assess the maternal mental health outcomes, random-effects meta-analysis was used to estimate the pooled effect sizes (ES) and its 95% for data on overall mean (SD) score for differences pre- and during pandemic. Furthermore, the overall pooled effect size was indicated by standardized mean differences (SMDs) before and during the pandemic. Cochrane’s Q and I-square statistics were applied to assess heterogeneity across included studies. I^2^ above 70% and Cochrane’s Q test with p < .05 indicated the presence of significant heterogeneity. Evidence for potential publication bias was examined using Egger’s and Begg’s tests.

### Quality and risk of bias assessment

Risk of bias was assessed separately for cross-sectional and non-cross-sectional study designs. For studies with an analytical cross-sectional design, we used the Joanna Briggs Institute (JBI) Critical Appraisal Checklist for Analytical Cross Sectional Studies. For studies with a longitudinal cohort, retrospective cohort, or case–control design, we used the appropriate version of the Newcastle–Ottawa Scale (NOS). In line with current guidance, we did not calculate a single summary score but instead examined patterns of strengths and limitations across domains for each study. We used these domain-level assessments qualitatively when interpreting the overall certainty of the evidence. Each item was judged as “yes”, “no”, or “unclear” based strictly on the information reported in the article. Where an item was not explicitly described, it was rated as unclear.

## Results

A total of 5,707 records were screened, of which 153 studies met the general inclusion criteria for the broader review. Among them, 23 studies met the more specific inclusion criteria and were included in this systematic review (Figure 1). These studies all assessed maternal mental health in women feeding their infants during the COVID-19 pandemic and provided a pre-pandemic comparator group using a validated mental health measurement tool.

**Figure 1.**
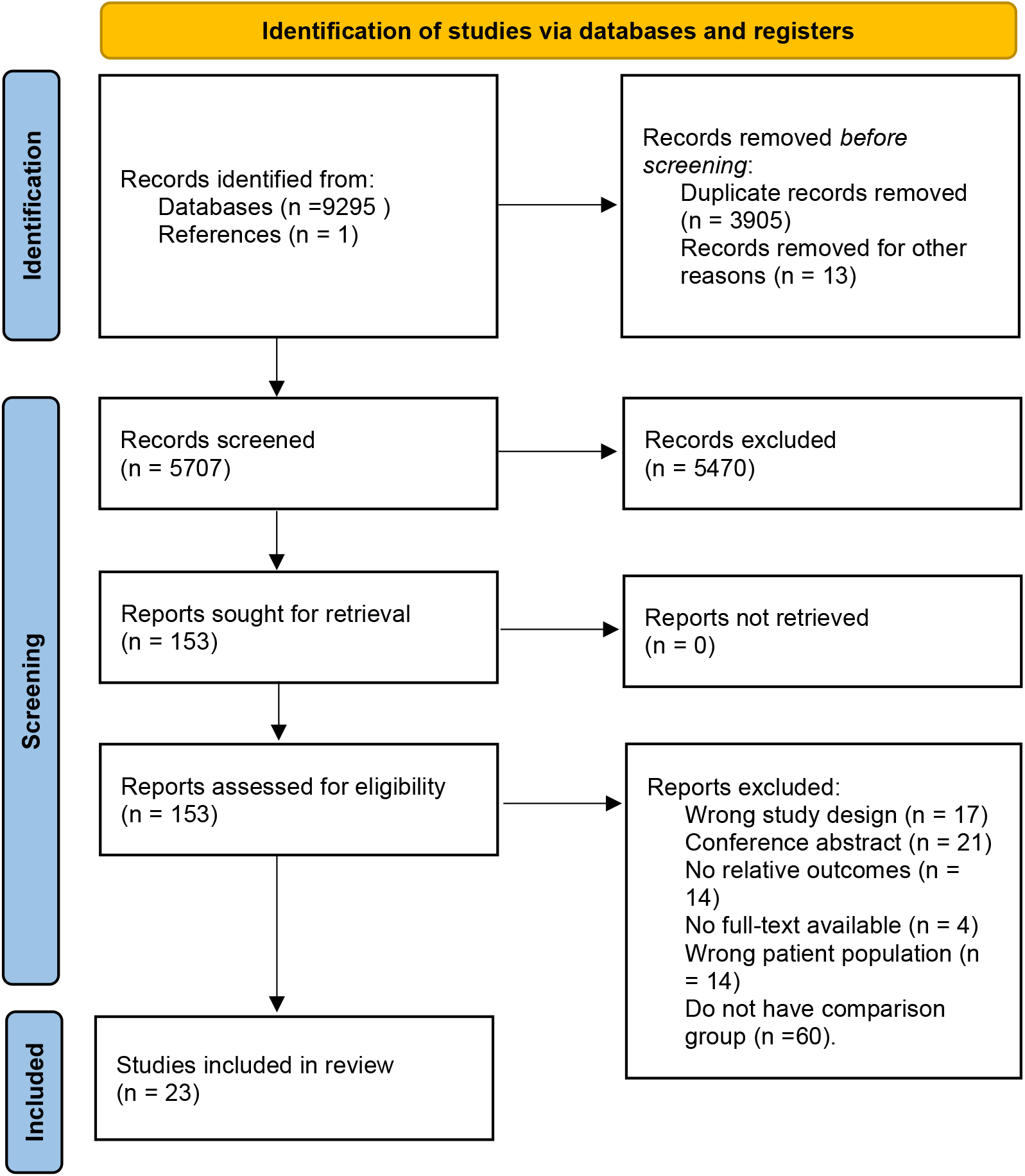
PRISMA flow chart of the review process. Source: Page MJ, et al. BMJ 2021;372:n71. doi: 10.1136/bmj.n71. This work is licensed under CC BY 4.0. To view a copy of this license, visit https://creativecommons.org/licenses/by/4.0/

### Study characteristics

The 23 included studies were conducted across 15 countries, with most originating from high-income settings such as the United Kingdom, Canada, the United States, Germany, and Japan. A minority of studies were conducted in middle-income countries, including Brazil and Turkey. The sample sizes ranged from 139 to 4,531 participants. Most studies were cross-sectional in design (n=18), while six used cohort or longitudinal designs. Detailed study characteristics are summarised in Table 1.

Maternal age was reported in 22 studies, typically ranging from 28 to 34 years. Educational attainment was inconsistently reported but generally indicated a high proportion of women with tertiary or university-level education. Delivery mode was reported in about two-thirds of the studies, with spontaneous vaginal delivery being the most common, while elective caesarean sections accounted for a smaller proportion. The infant age at the time of study varied widely across studies, ranging from birth to 12 months, with Zanardo et al [10] and Pariente et al assessing mothers within 2 days after delivery. Eight studies did not report infant age.

Where possible, outcomes were synthesised quantitatively using meta-analysis. However, several studies reported mental health outcomes as proportions above predefined cut-off scores rather than as means with measures of dispersion. These studies could not be included in meta-analyses and were therefore summarised descriptively.

### Mental health outcomes

All included studies used at least one validated tool to assess maternal mental health. The most commonly used instrument was the EPDS, reported in 16 studies, particularly in studies from Europe and North America. Other tools included the Generalized Anxiety Disorder Scale (GAD-7, n=5), State-Trait Anxiety Inventory (STAI, n=3), Perceived Stress Scale (PSS, n=3), Patients Health Questionnaire-2 (PHQ-2, n=2), and some questionnaires identified in one study each: Beck Depression Inventory (BDI), PTSD checklist, validation study of the Hospital Anxiety and Depression Scale (HADS), and the Parental Stress Scale. Eleven studies used more than one mental health instrument.

### Maternal depression assessed by EPDS

Eleven of the 16 studies that reported EPDS scores provided mean (SD) value for pre-COVID and COVID periods. Pooled analysis showed a small but statistically significant increase in depressive symptoms among mothers during the pandemic (SMD = 0.21, 95% CI: 0.14 to 0.29). The direction of effect was consistent across most studies, apart from Yakupova et al. [11]. The forest plot and individual study effect sizes are presented in Figure 2.

**Figure 2.**
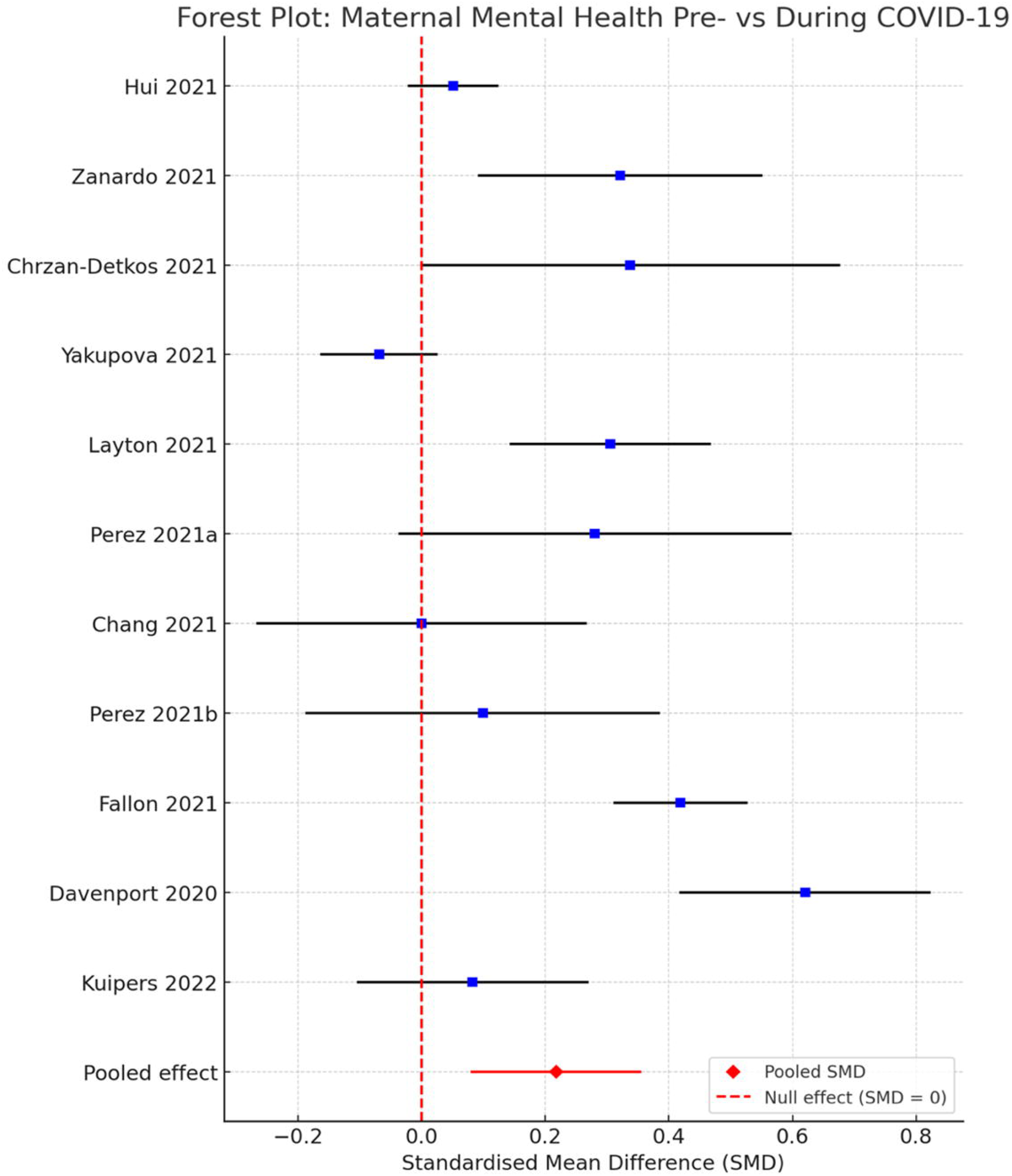
Forest plot of maternal postnatal depression assessed by EPDS.

### Maternal anxiety by GAD-7 or STAI

Three of the five studies that measured anxiety used the GAD-7 were eligible for meta-analysis(Figure 3). The pooled standardised mean difference indicated a significant increase in maternal anxiety during the COVID-19 pandemic compared to the pre-pandemic period (SMD = 0.27, 95% CI: 0.13 to 0.41).

**Figure 3.**
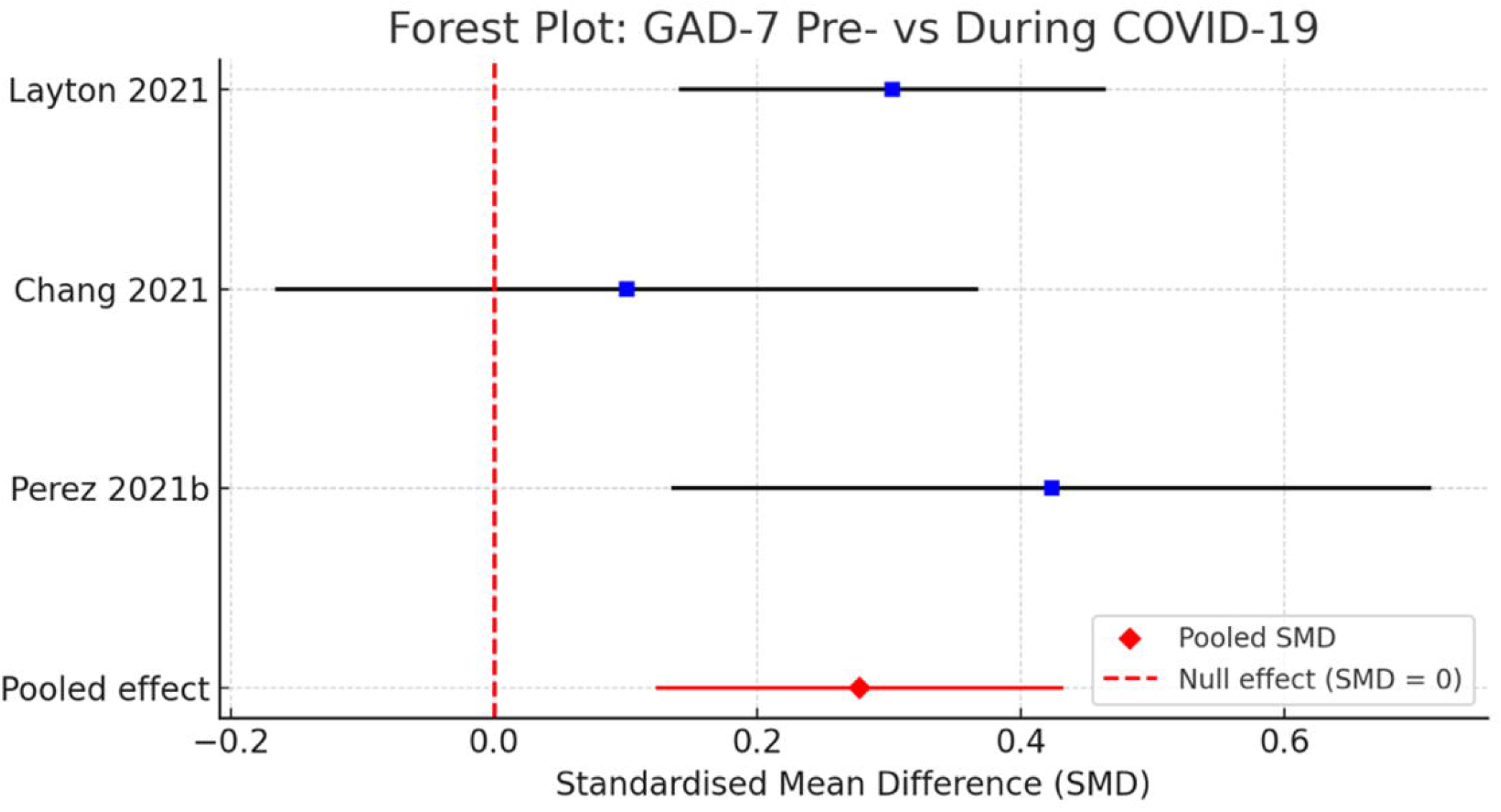
Forest plot of maternal postnatal anxiety assessed by GAD-7.

For STAI, a pooled analysis of two studies also suggested an increase in maternal state anxiety (pooled SMD = 0.35). However, the confidence interval indicated marginal statistical significance and substantial between-study heterogeneity (I^2^ = 88.0% and 88.6% for EPDS and STAI separately, whilst the GAD-7 analysis showed low-to-moderate heterogeneity I^2^ = 28.0%).

### Maternal stress by PSS

Three studies assessed maternal stress during the COVID-19 pandemic using the PSS. However, only two studies provided comparable summary statistics and due to the heterogeneity in outcome reporting, a meta-analysis was not performed for this outcome and findings were synthesised descriptively.

Janevic et al. conducted a survey in USA, where mothers who gave birth after the COVID-19 period reported varied stress levels [70.4% scored in the “less stressed” range (PSS≤20) vs. 29.6% in the “more stressed” range (PSS≥21)]. Comparatively, a large Dutch cohort study by Juncker et al. included 2310 women delivering during the pandemic and 151 pre-pandemic controls. Results showed non-significantly higher mean PSS scores during the pandemic (19.56 ± 7.97) compared with the pre-pandemic group (18.69 ± 0.47). Similarly, a Polish cross-sectional survey by Kolomanska-Bogucka et al. reported no significant increase in perceived stress between the pre-pandemic group (median PSS: 18.0, IQR 13.0–22.0) and two COVID-19 cohorts (17.0, IQR 13.0–22.0 in both).

### Other maternal mental health outcomes

In addition to depression, anxiety, and stress, several studies examined parental stress and bonding. Fernandes et al. [12] used the Parental Stress Scale (PSS) and the Postpartum Bonding Questionnaire (PBQ) in a Portuguese cohort and found that while 27.5% of mothers showed clinically significant depressive or anxiety symptoms during the pandemic, there were no significant differences in mother–infant bonding scores compared with pre-pandemic levels. Babu et al. [13] reported a large study conducted in the US (n= 2205 during pandemic; n = 540 pre-pandemic) with evaluation of posttraumatic psychological growth (PTG; a self-report measure used to assess the positive psychological changes an individual experiences after a traumatic event), maternal acute stress responses, and bonding. They found that approximately 60% of mothers reported experiencing some degree of PTG following childbirth. Among those delivering during the pandemic, higher levels of acute childbirth-related stress were associated with greater PTG (β = 0.07, p<0.01). In turn, higher PTG scores were linked to better mother–infant bonding (β = 0.22, p<0.001) and fewer posttraumatic stress symptoms (β = -0.06, p<0.05). These indirect effects via PTG were only significant for mothers delivering during COVID-19 and not for those delivering before the pandemic.

Additionally, a retrospective cohort study from an urban US hospital evaluated the impact of telemedicine on postpartum care during the pandemic [14]. While overall postpartum visit attendance rates remained similar across pre-COVID, peak-COVID, and ongoing-COVID periods (52%, 43%, and 56%, respectively; p>0.05), there was a significant reduction in postpartum depression screening rates during the pandemic (22% during peak-COVID and 33% ongoing-COVID vs. 74% pre-COVID, p<0.01). No significant differences were observed in hospital readmissions, contraceptive use, or breastfeeding rates in this study.

### Quality of the included studies

Eighteen studies were appraised with the JBI analytical cross-sectional checklist. As shown in Figure 5, Seventeen of 18 studies clearly defined their inclusion criteria (item 1). Apart from Davenport et al. [15], all studies described the study population and setting in sufficient detail (items 2), measured the exposure (pre-pandemic versus pandemic period) using objective calendar dates (item 3), and used standardised criteria for mental health outcomes (item 4). Most studies used valid and reliable outcome measurement (item 7), except Yakupova et al. [11], Perez et al.[16], and Davenport et al. [15]. Seven of 18 studies explicitly identified potential confounding factors, such as maternal age, education, parity, socioeconomic status or previous mental health problems (item 5), and the same seven studies reported clear strategies to address confounding, typically through multivariable regression models (item 6); sensitivity analysis showed no significant differences between the adjusted studies and those with no adjustment (SMD = 0.14 (95% CI 0.01, 0.27) vs. SMD = 0.20 (95% CI 0.06, 0.35). Five studies [15,17-20] did not mention confounders at all, and six studies [12-14,21-23] provided incomplete information with unclear risks. All cross-sectional studies were judged to have used appropriate statistical analyses for their design and research questions (item 8).

**Figure 4.**
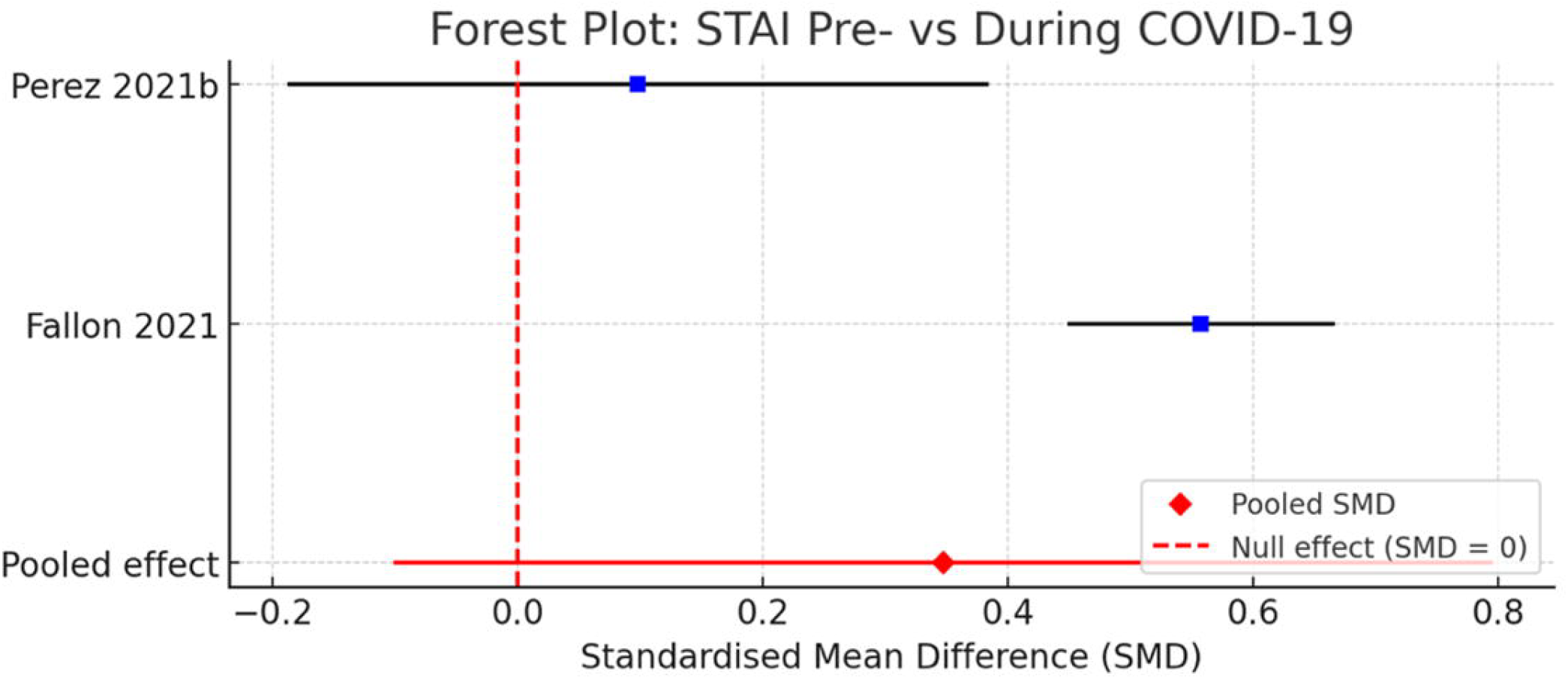
Forest plot of maternal postnatal anxiety assessed by STAI.

**Figure 5.**
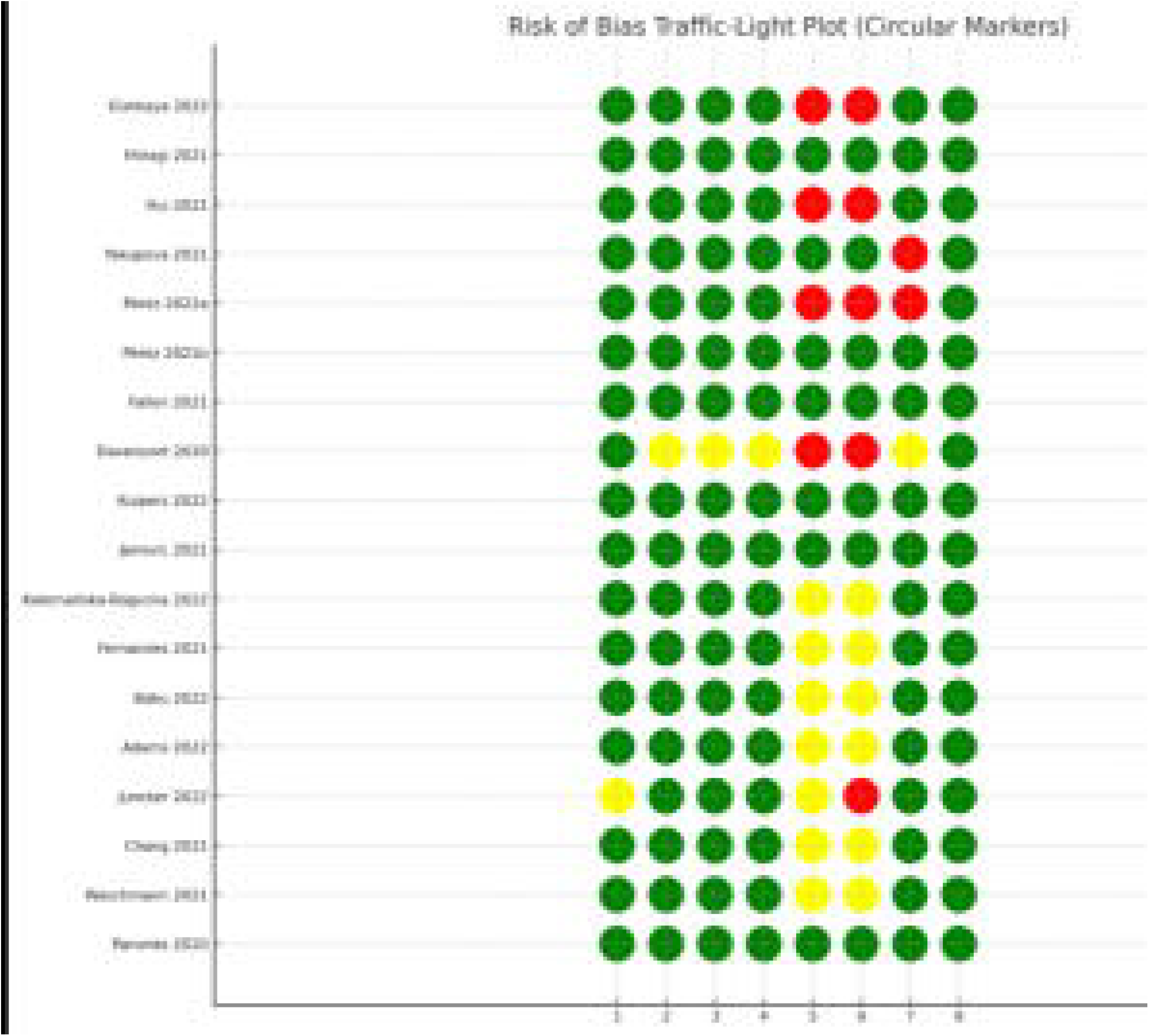
Risk-of-bias plot of the included studies Notes: The checklist includes eight items (domains): (1) clearly defined inclusion criteria, (2) description of study subjects and setting, (3) valid and reliable measurement of the exposure, (4) use of objective, standard criteria for outcome measurement, (5) identification of potential confounding factors, (6) strategies to deal with confounding factors; sensitivity analysis showed no significant differences between the adjusted studies and those with no adjustment (SMD = 0.14 (95% CI 0.01, 0.27) vs. SMD = 0.20 (95% CI 0.06, 0.35). (7) valid and reliable outcome measurement, and (8) appropriateness of the statistical analysis. Each item was judged as “yes” (Green), “no” (Red), or “unclear” (Yellow) based strictly on the information reported in the article. Where an item was not explicitly described, it was rated as unclear.

Two cohort [24,25] and one case–control [10] studies were appraised using the NOS. In general, these studies had adequate selection of exposed and non-exposed groups, and exposure and outcome were ascertained from reliable sources (hospital records or validated questionnaires). Zanardo et al. 2021 [10] comparing births during lockdown with pre-pandemic births used the same hospital setting and identical outcome measurement in cases and controls, but did not exclude women with depressive symptoms from the control group and did not clearly control for potential confounders in the comparative analyses.

## Discussion

This systematic review and meta-analysis examined postpartum depression, anxiety, and stress among breastfeeding mothers during the COVID-19 pandemic. Across 23 studies, we found consistent evidence that depressive and anxiety symptoms increased during the pandemic compared with pre-pandemic periods, based on validated tools including the EPDS, GAD-7 and STAI, suggesting that a large proportion of breastfeeding mothers experienced psychological difficulties during the pandemic worldwide.

Our meta-analysis showed a small increase in depressive symptoms measured by EPDS during the pandemic (SMD=0.21). Although this effect size cannot be directly translated into clinical thresholds, it suggests a potential shift in symptom burden at the population level. Previous reviews of perinatal mental health during COVID-19 have consistently reported increased levels of depression and anxiety across different countries [26-28]. These studies mainly using pooled prevalence estimates rather than continuous symptoms scores; however, our results are consistent with those findings and provide a specific pooled effect size. A global meta-analysis reported a pooled depression prevalence of 34% in pregnant and postpartum women, which was substantially higher than pre-pandemic estimates [29]. Compared to perinatal mothers, postpartum breastfeeding mothers may have been particularly affected during Covid-19 lockdowns, because breastfeeding benefits from regular contact with healthcare professionals and lactation support, which was heavily disrupted in many countries [30]. Moreover, studies also showed that mothers of young infants experienced particularly high levels of loneliness, psychological distress, and fear of infection during the pandemic [5]. Studies from the UK [31], US [32] and Australia [33] found that job loss, reduced income, or unstable childcare arrangements were also strongly associated with worse maternal mental health during the pandemic. These issues may have been intensified for breastfeeding mothers, who may be more reliant on in-person professional and peer support during the early postpartum period [6].

Our review also found increased anxiety symptoms during the pandemic. The pooled effect for GAD-7 (SMD=0.27) closely matches results from general population-based studies. One of our included studies conducted in the UK showed that postpartum anxiety scores measured by STAI nearly doubled during the first lockdown [15]. A Canadian longitudinal study also reported notable increases in anxiety and stress among postpartum women during 2020 compared with earlier years [6]. Breastfeeding mothers may have faced additional pressures, including concerns about virus transmission, the safety of feeding practices [34], and reduced support from partners, family, and health professionals due to restrictions [35,36].

Comparatively, measurements of stress during the pandemic were not significantly different from those pre-pandemic. Two studies using the PSS [20,21] reported no clear differences between pre- and during-pandemic periods, whilst two studies using other measurements of stress [12,13] showed a non-significant increase. Stress responses likely varied depending on timing, national mitigation policies, financial stability, and access to health and community services [6,37,38]. Differences between countries may also reflect variations in social protection systems. For example, Juncker et al. 2022 [20] reported relatively stable stress levels in mothers in the Netherlands, which could reflect consistent access to their local healthcare and employment support during the pandemic period; this was also mentioned in a recent preprint of another Dutch study [39].

Several included studies also examined parental stress, bonding, and post-traumatic growth during the pandemic. Although depressive and anxiety symptoms increased, mother-infant bonding did not appear to decrease in the limited studies that measured it [12,13,25]. One US study found that childbirth-related stress could even relate to greater post-traumatic growth measured by Posttraumatic Growth Inventory, which in turn was linked to better mother-infant bonding [13]. This suggests that some mothers adapted positively despite challenging circumstances. However, these findings should be interpreted with caution as they were measured by different approaches and were not consistent across all studies.

This review has several limitations. First, most included studies were cross-sectional, limiting our ability to infer causality. Differences between pre- and during-pandemic groups may reflect unmeasured confounding rather than the impact of the pandemic itself. Second, many studies lacked detailed reporting of socioeconomic characteristics or previous mental health conditions, making residual confounding likely. However, the direction of findings was consistent across studies with both low and moderate risk of bias. Therefore, the observed increase in postpartum depression and anxiety may not be solely due to bias or methodological differences. Third, heterogeneity was high across outcomes, reflecting differences in timing, measurement tools, pandemic phases, and healthcare contexts. Fourth, some studies relied on convenience samples or online recruitment, which may limit generalisability. Finally, we were unable to examine subgroup differences by breastfeeding exclusivity, parity, ethnicity, or socioeconomic status due to limited reporting.

A strength of this systematic review and meta-analysis is its focus on breastfeeding mothers, a group often overlooked in broader perinatal mental health research. Our findings provide insights into how this vulnerable group responded to the pandemic crisis and provide evidence relevant for policymakers about how to protect maternal wellbeing in future public health emergencies. Although COVID-19 has passed, we should learn from this experience. Health systems should keep breastfeeding support available, make sure mothers can access mental health screening, and allow flexible service models such as telehealth and community support. These steps can help health systems stay prepared and resilient in any potential public emergencies in the future.

## Data Availability

All data produced in the present study are available upon reasonable request to the authors

## Declaration

The authors declare that they have no competing interests.

