## Supplementary material for "Pandemic-related changes in postpartum depression and anxiety among breastfeeding mothers: a systematic review and meta-analysis": Table 1

Table 1. Characteristics of the included studies.

| **Study ID** | **Country** | **Study design** | **Total number of participants** | **Maternal age (years)** | **Maternal education** | **Delivery mode** | **Infant age at time of study** | **Infant gestational age** | **Validated tools** | **Mental health outcomes** | **Other maternal related results** |
| --- | --- | --- | --- | --- | --- | --- | --- | --- | --- | --- | --- |
| Fiske 2022 | UK | longitudinal cohorts | Study 1 n=175 (84 with both visits before pandemic, 70 with second visit during pandemic, 21 with both visits during pandemic)  Study 2 n=220 (154 also in Study 1) | not report | not report | Not report | not report | not report | BDI | BDI-II: Pre-Covid: T1 (10 months): 10.64 (SD 9.31) ; T2 (16 months) 9.90 (SD 10.12) Pre to During: T1 (10 months) 9.54 (SD 9); T2 8.57 (SD 8.77) | Maternal Stress over Pandemic: Decrease in stress, increase in pandemic-specific depression over six months. |
| Gunkaya 2022 | Turkey | cross-sectional | 210 total (98 April, 106 July) | 28.15 (SD 5.7) | not report | Not report | not report | not report | EPDS | Overall: 3.93 (SD 4.85) April: 4.38 (SD 5.21) July: 3.51 (SD 4.48) | EPDS Scores Over Time: EPDS and PSQI scores remained similar; BSES scores improved significantly. |
| Hiiragi 2021 | Japan | Cross-sectional, retrospective, in-person questionnaire | Pre-pandemic n=339, pandemic n=279 | Pre-pandemic median 33, pandemic median 33 | not report | Not specified - 'maternal postpartum complications' rate was 4.4% pre-pandemic and 1.8% during pandemic | 28-35 days as defined in inclusion criteria | Not specified but preterm % was 9.1 pre-pandemic and 11 during pandemic | EPDS | EPDS>9 Pre-Covid: 34 (10%) Covid: 38 (14%) | EPDS Scores Pre vs. Pandemic: No significant difference in EPDS scores or behavioural changes. |
| Hui 2021 | China, Hong Kong | Cross-sectional | 4531 (total): 3577 (the pre-alert group); 954 (post-alert group) | pre-alert group: 33.1 (SD=4.4) post-alert group: 33.1 (SD=4.6) | not report | Pre-alert group: Normal vaginal: 2175 (60.8%); Instrumental delivery: 378 (10.6%); C-section: 1005 (28.1%).  Post-alert group: Normal vaginal: 594 (62.3%); Instrumental delivery: 91 (9.5%); C-section: 262 (27.5%) | not report | Pre-alert group:38.5 ± 2.25; Post-alert group: 38.5 ± 2.29 | EPDS | EPDS 1-day after delivery Pre-alert: 4.71 (95%CI 4.57-4.85) Post-alert: 4.93 (95%CI 4.66-5.21) | EPDS Post-Delivery: 96.2% scored day after, 83.2% within a week; higher in post-alert group. |
| Zanardo 2021 | Italy | Non-concurrent case-control | Pre-pandemic n=147 Pandemic n=152 | Pre-pandemic: mean 33.18, SD 5.10 Pandemic: mean 33.47, SD 4.93 P value 0.618 | Pre-pandemic: 36.73% degree Pandemic: 44.08% degree P value 0.238 | Pre-pandemic: 12.93% C-section Pandemic: 17.76% C-section P value 0.265 | Data collected at 2nd day postpartum | Pre-pandemic: mean 39.73 weeks, SD 1.13 Pandemic: mean 39.68 weeks, SD 1.19 P value 0.713 | EPDS | During Covid: Total EPDS: 8.03 (SD:4.88); >12 EPDS n=35 (23.03%)  Before Covid: Total EPDS: 6.58 (SD:4.08); >12 EPDS n=35 (23.03%)n=17 (11.56%) | EPDS Scores: Pre vs. Pandemic: Significant increase in pandemic EPDS scores (8.03) and anhedonia. |
| Chrzan-Detkos 2021 | Poland | Retrospective, observational | Total n=139. Pre-pandemic group n=61. Pandemic group n=78. | Pre-pandemic group: Mean age 31.04, SD 3.70 Pandemic group: Mean age 31.74, SD 5.06 | Not significantly different between 2 groups - mean would be closer to higher education than secondary education but data presented in confusing manner. | Vaginal: Pre-Covid 59.02% Covid 53.84% | not report | Pre-Covid: 38.56 (SD 3.55) Covid: 38.88 (SD 1.88) | EPDS | EPDS: Pre-Covid: 13.56 (SD 6.46) Covid: 15.71 (SD 6.23) | Shift in Location and PPD Symptoms: Move from large to medium cities; increased PPD severity during pandemic. |
| Pariente 2020 | Israel | Cohort study, comparison with previous data from same medical centre, in-person questionnaire day 2 post-partum | Pandemic group n=223 Pre-pandemic group n=123 | Pandemic mean 29.1, SD 5.1 Pre-pandemic mean 28.3, SD 5.0 | not report | not report | Questionnaire distributed at 2 days post-partum | Pandemic 39.4 weeks, SD 1.0 Pre-pandemic 39.4 weeks, SD 0.9 | EPDS | EPDS>13 :Before Covid:15.2%; During Covid: 6.8%. EPDS>10: Before Covid:31.3%;  During Covid:16.7% | Lower Rate of PPD During Pandemic: Surprisingly, a statistically significant lower rate of PPD during the pandemic. |
| Waschmann 2022 | USA | Retrospective chart review | Total n=1061 Pre-pandemic n=557 Pandemic n=504 | Pre-pandemic: mean 31.8, SD 5.33 Pandemic: mean 31.4, SD 5.48 P value 0.28 | not report | Pre-pandemic: 74.1% vaginal Pandemic: 73.0% vaginal P value 0.73 | Not specified - as per inclusion criteria had to have at least one valid EPDS score within 3 months postpartum | Pre-pandemic: mean 39.3 weeks, SD 1.16 Pandemic: mean 39.3 weeks, SD 1.12 P value 0.64 | EPDS | EPDS >10: pre-Covid:16.9%; During Covid: 18.1% | PPD Incidence in 2019 vs. 2020 Epochs: Similar PPD incidence (16.9% to 18.1%); no significant differences. |
| Yakupova 2021 | Russia | Cross-sectional, online questionnaire | Pre-pandemic group n=611 Pandemic group n=1645 | Pre-pandemic mean 31.17, SD 4.54 Pandemic mean 30.98, SD 4.42 | Pre-pandemic tertiary/university 90.7% Pandemic tertiary/university 91.8% | Pre-pandemic 77/4% vaginal Pandemic 71.9% vaginal | Pre-pandemic 6.37 (SD 3.42) Pandemic 6.93 (SD 3.30) | Pre-pandemic 39.47 weeks (SD 1.67) Pandemic 39.40 weeks (SD 2.04) | EPDS CBTS | EPDS: Before Covid:9.88 (SD6.07); During Covid: 9.46 (SD 6.13), p=0.15. CBTS:Before Covid:17.16 (SD: 11.35); During Covid: 15.83 (SD 11.40),p=0.014 | Pre vs. Pandemic EPDS/CBTS Scores: Pre-pandemic EPDS 9.88, Pandemic 9.46; not significant. CBTS showed significant decrease during pandemic. |
| Layton 2021 | Canada | Cross-sectional, using data from pre-existing RCTs. | Pre-pandemic n=305 Pandemic n=298 | Pre-pandemic mean 32.94, SD 5.17 Pandemic mean 31.74, SD 4.78 | Pre-pandemic mean 16.94 years of education Pandemic mean 16.76 years | not report | Pre-pandemic mean 6.04 months, SD 4.18 Pandemic mean 5.32 months, SD 3.66 | not report | EPDS GAD-7 | **Pre-covid group** (p<0.01) EPDS: 14.93 (SD 4.44) GAD-7: 10.88 (SD 4.93) **Covid group** (p<0.01) EPDS: 16.28(SD 4.39) GAD-7: 12.39 (SD 5.05) | EPDS and GAD-7 Scores Pre vs. Pandemic: Significant increase in pandemic EPDS (16.28) and GAD-7 (12.39) scores. |
| Perez 2021a | Germany | Prospective, cross-sectional, using data from larger longitudinal study | Control n=97 Covid n=65 | Control: mean 34.51, SD 3.25 Covid: mean 36.02, SD 4.55 | Control: 70.1% university degree Covid: 78.5% university degree | not report | Study conducted at 7 months postpartum | not report | EPDS | EPDS: Before Covid:5.05 (SD:4.37); During Covid: 6.25 (SD 4.08), p=0.046. | EPDS Scores Pre-COVID vs. COVID: Increased mean EPDS score during COVID; statistically significant. |
| Chang 2021 | Canada | Observational a | Total n=220 Pre-covid cohort n=100 Covid cohort n=120 | Mean age in pre-covid cohort = 31.4 (SD 4.7) Mean age in covid cohort = 32.0 (SD 4.9) | Years of education:  Pre-Covid: 14.9 (SD 1.8) Covid: 15.2 (SD 1.7) | not report | Not specified - inclusion criteria stated infant <12 months | not report | EPDS GAD-7 | EPDS: Pre-Covid: 16 (SD 4), Covid 16 (SD 4.7) GAD-7:  Pre-Covid: 12.4 (SD 4.5), Covid: 12.9 (SD 5.3) | Comparative Study Pre/Post-COVID: No significant differences in EPDS, PSWQ, GAD-7, DAS, or SPS scores. |
| Perez 2021b | Germany | Single-centre, cross-sectional, using control data from pre-existing longitudinal study (PAULINE study) | Control n=101 Covid n=90 | Control: mean 33.7, SD 4.08 Covid mean 34.9, SD 4.54 | Control: 69.0% university degree Covid: 76.7% university degree | Control: 78.2% spontaneous vaginal delivery Covid: 58.9% spontaneous vaginal delivery | Control: mean 22.42 days, SD 6.72 Covid: mean 54.56 days, SD 9.77 P value <0.001 | Not specified (premature births <37 weeks were excluded) | EPDS STAI GAD-7 | EPDS: Before Covid:6.24 (SD:4.36); During Covid: 6.68 (SD 4.51), p=0.50. STAI:Before Covid:33.3 (SD: 6.27); During Covid: 33.9 (SD 5.89),p=0.52. GAD-7:Before Covid:3.63 (SD:2.58); During Covid: 4.75 (SD: 2.7), p=0.01 | Birth Experience During COVID-19: No significant differences in depression, anxiety, social support, or birth experience. |
| Fallon 2021 | UK | Cross-sectional, online survey | n=614 | Mean 30.9, SD 5.1 | Postgraduate education 24.4%, undergraduate 40.4% | 51.5% uncomplicated vaginal, 11.9% assisted vaginal, 18.4% elective C-section, 18.2% emergency C-section | Mean weeks 7.0, SD 3.6 | 7.4% premature (<37 weeks), 0.5% post term (>42 weeks) | EPDS STAI PSAS | EPDS:  Pre-Covid:9.13 (SD 5.72) Covid:11.56 (SD 5.9) STAI:  Pre-Covid: 37.7 (SD 13.45) Covid:45.26 (SD 13.69) PSAS:  Covid: 24.79 (SD 6.19) | EPDS and STAI Scores During Pandemic: Significant rise in EPDS and STAI scores; increased motherhood-related anxiety. |
| Davenport 2020 | Multinational (online survey) - 72.8% Canadian, 8.1% British, 5.9% USA | Cross-sectional, multinational, online | Total n=900 Pregnant n=520 First year after delivery n=380 | Median age 33, range 17-49 | Some form of post-secondary education n=520 | not report | not report | not report | EPDS  STAI | EPDS: Pre-Covid: 7.5 (SD 4.9) Covid: 11.2(SD 6.3) STAI Pre-Covid: 34.5 (SD 11.4) Covid: 48.1 (SD 13.6) | Maternal Depression and Anxiety Increase: Significant increase in depression (40.7%) and anxiety (72%) during pandemic. |
| deMola 2021 | Brazil | Cohort | 1142 | 27.5 (SD 6.5) | not report | Not report | 11.4 (SD3.7) | not report | EPDS, GAD-7, IES | EPDS: Pre-Covid: 5.1%  Covid: 29.5% GAD-7: Pre-Covid: 9.7% Covid: 25.9%  IES (moderate/severe): Covid: 40.6% | Stress and Mental Health During Pandemic: High rates of stress, depression, and GAD; significant increases from baseline. |
| Kuipers 2022 | Belgium | Cross-sectional, online questionnaire | Total postpartum n=604 Pre-pandemic group n=456 Pandemic group n=148 | Pre-pandemic group: 30.53 Pandemic group: 30.44 | Pre-pandemic group: 'high level' 79.2% Pandemic group: 'high level' 81.1% | Pre-pandemic group: spontaneous vaginal 62.7% Pandemic group: no data ?reason | Pre-pandemic group: 23.76 weeks Pandemic group: 16.66 weeks | not report | EPDS GAD-2 | **Total** EPDS:8.01 (SD 4.82) GAD-2: 2.18 (SD 1.47) **Pre-covid group** EPDS: 7.76 (SD 4.86) GAD-2: 1.98 (SD 1.33) **Covid group** EPDS: 8.16 (SD 4.79) GAD-2: 2.29 (SD 1.53) | GAD-2 Scores Pre vs. During Pandemic: Significant differences; small positive effect of having an infant during COVID-19. |
| Janevic 2021 | USA | cross-sectional | delivered prior to March 15 (n=654); delivered after March 15 (n=1441) | 19-24: 7(3%); 25-29:27(11.6%); 30-34:90 (38.8%); 35-39: 85 (36.6%); 40-49: 23 (9.9%) | not college degree: 37 (15.7%) college degree: 198 (84.3%) | not cesarean: 118 (69.4%); cesarean: 52 (30.6%) | not report | term: 158 (93.5%); pre-term: 11 (6.5%). | GAD-7; PSS:PHQ-2 | GAD-7  less anxious (0-9):n=191 more anxious (10-21) n=37  PSS less stressed (0-20) n=157 more stressed (21-40) n=66  PHQ-2 not depressed (0-2) n=210 depressed(3-6) n=13 | Birth Satisfaction and PTSD: Higher satisfaction lowers anxiety, PTSD; healthcare discrimination increases both. |
| Fernandes 2021 | Portugal | Cross-sectional online study | 567 (414 pre-pandemic, 153 during pandemic) | pre-Covid 32.95, covid 33.02 | Basic: precovid 44.9%, covid 41.2% Higher: precovid 55.1%, covid 58.8% | not report | Precovid 6.23 (2.81)m, covid 1.34 (0.75)m | not report | HADS, Parental Stress Scale | Depression (HADS):  Pre-Covid: 6.03 (SD 3.86) Covid:5.50 (SD 3.76)  Parenting Stress (PSS) Pre-Covid: 35.96 (SD 8.38) Covid:37.40 (SD 9.03) | Mothers' Anxious/Depressive Symptomatology: 72.5% normal or mild; 27.5% clinically significant levels. |
| Babu 2022 | USA | Cross-sectional | 2205 during pandemic, 544 Pre-pandemic | 31.98 (SD 4.54) | Bachelor's degree (78.1 %) | Vaginal (71%) | 2.23 (SD 1.53). | Term (93.3 %) | PDI PTSD | Posttraumatic psychological growth (PTG) significantly different between before and during pandemic (X2=66.56, p=0.0001, PTSD as covariate). Acute stress response significantly different between before and during pandemic (X2=23.72, p=0.0001, PTG as covariate) | Acute stress linked to elevated PTG, reducing posttraumatic stress symptoms and improving bonding. |
| Adams 2022 | USA | Retrospective cohort study | 359 (142 pre-Covid, 133 peak-Covid, 85 ongoing-Covid) | Pre 29.5 peak-Covid 29.4 ongoing-Covid 30.3 | not report | CS pre 35.2%, peak 34.1%, ongoing 32.9% | not report | 38 all groups | PHQ2 | PHQ-2: Pre 74%, Peak 22%, Ongoing 33% | Prenatal Visit and Telemedicine Usage: Fewer prenatal visits during ongoing-COVID; significant increase in telemedicine postpartum care. |
| Juncker 2022 | Netherlands | cohort | Covid=2310 (covid positive n=691) pre-covid=151 | 33.1 (SD 3.8) | Bachelor equivalent 44.5%, masters and doctoral equivalent 37.2% | 82.1% vaginal, 5.8% instrumental, 12% C-section | mean 34.0 weeks | mean 40.0 weeks | PSS | PSS 19.56 (SD 7.97), Pre-covid : 18.69 (SD 0.47) | PSS Scores and COVID Antibodies: No significant increase in PSS scores; no correlation with COVID antibodies in breast milk. |
| Kolomanska-Bogucka 2022 | Poland | Cross-sectional, in-person questionnaire | Pre-pandemic group: n=252 COVID-1 group (before Sep 2020): n=262 COVID-2 group: n=226 | Pre-pandemic group: 31.0 COVID-1 group: 31.0 COVID-2 group: 31.0 | Pre-pandemic group: 80.9% university COVID-1 group: 76.7% university COVID-2 group: 81.4% university | Pre-pandemic group: 63.7% vaginal COVID-1 group: 58.0% vaginal COVID-2 group: 54.0% vaginal | not report | Pre-pandemic group: delivery at term 59.8% COVID-1 group: delivery at term 61.8% COVID-2 group: delivery at term 55.8% | PSS | **Pre-pandemic group**: Median PSS 18.0 (Q1 13.0, Q3 22.0) **COVID-1 group**: 17.0 (13.0, 22.0) **COVID-2 group:** 17.0 (13.0, 22.0) | Physical Activity Levels During Pandemic: Reduced energy in physical activities; significant differences between groups. |

Notes: Data are presented as mean (standard deviation, SD), median (interquartile range, IQR), percentage, or number of participants unless otherwise stated. Mental health outcomes were assessed using validated instruments including the Edinburgh Postnatal Depression Scale (EPDS), Generalized Anxiety Disorder Scale (GAD-7), State-Trait Anxiety Inventory (STAI), Beck Depression Inventory (BDI), Perceived Stress Scale (PSS), Hospital Anxiety and Depression Scale (HADS), Postpartum Bonding Questionnaire (PBQ), and other validated questionnaires as reported in the original studies. “Pre-pandemic” refers to participants assessed before the COVID-19 pandemic or before implementation of pandemic-related restrictions, while “pandemic” refers to participants assessed during the COVID-19 pandemic period.
